# Health effects of European colonization: An investigation of skeletal remains from 19th to early 20th century migrant settlers in South Australia

**DOI:** 10.1101/2021.04.18.21255521

**Authors:** Angela Gurr, Jaliya Kumaratilake, Alan Henry Brook, Stella Ioannou, F. Donald Pate, Maciej Henneberg

## Abstract

Nineteenth century medical understanding of human metabolism was limited, therefore, the incidence of metabolic deficiencies was not fully recorded. In addition, the transition from agricultural based mode of life to the industrial one significantly changed the pattern of these metabolic deficiencies. They were further altered by colonisation of distant continents. Palaeopathological study of skeletal remains from the early industrialised colonial era allowed light to be shed on the metabolic stresses produced by this new mode of life. Aims of this study were to investigate manifestations of disease in skeletal remains from 65 (20 adults, 45 sub-adults) migrant settlers buried in the “free ground” of St Mary’s Anglican Church Cemetery (1847 to 1927). An area allocated for burials paid for by the South Australian Government. Skeletal manifestations were determined and interpreted in terms of their multiple aetiologies. Findings were compared with those published for two 19^th^ century British samples. Skeletal manifestations, commonly related to metabolic deficiencies, were observed. Areas of abnormal porosity of bone cortices were seen in 9 adults and 12 sub-adults, flaring of metaphyses was seen in one sub-adult, flaring of costochondral junctions of the ribs was seen in one sub-adult. Porous lesions of orbital roof bones (Types 3 to 5) were seen on three sub-adults. Micro-CT scans of tooth samples located interglobular dentine in three individuals. Comparison of St Mary’s findings with St Martins, Birmingham, and St Peter’s, Wolverhampton, UK, showed more individuals from St Mary’s had areas of abnormal porosity of bone cortices possibly related to vitamin C deficiency. However, St Mary’s sample displayed fewer changes attributable to vitamin D deficiency as expected in a country with greater UV irradiation. This indicates that, although the early industrialisation produced metabolic stresses, change of the environment through colonisation of new continents altered the distribution of metabolic deficiencies.

## Introduction

Although medical knowledge and medical literature had been well developed in the 19^th^ century, they did not include the modern understanding of human metabolism and thus could not record the occurrence of metabolic deficiencies. Pattern of these deficiencies had been significantly changed by the transition from agricultural based mode of life to the industrial one and further altered by colonisation of far away continents. Palaeopathological study of skeletal remains from the early industrialisation colonial era allows to shed light on the metabolic stresses produced by the new mode of life.

Industrialisation of Britain in the 19^th^ century altered the landscape, the economy, and the lives of many people. This had health consequences. The expansion of urban centres of manufacture offered opportunities for employment, however, the majority of cities lacked the infrastructure to cope with the increase in the population size due to a migration of workers. Many people had to live under overcrowded and unsanitary conditions, where the water supplies could be contaminated [1-3]. These conditions, long hours of working inside factories powered by coal fired engines, and diets lacking in essential nutrients contributed to the poor health of the working classes in a way different from traditional agricultural communities [2, 4-7].

Rapid development of urban centres, also affected the traditional small industries in rural regions of Britain.The inability to compete with the low production costs of commercial items in large factories caused an increase in rural unemployement [8-10]. In addition, importation of large quantities of raw materials to feed the urban industries at cheaper prices, such as iron, copper and tin ores, caused the down turn of regional mining industries [9, 10]. Poor harvests and the rapid spread of potato blight, further affected farmers [8, 11]. The net outcome was economic hardship in regional rural areas of the United Kingdom.

The British Government planned to reduce over-crowding in industrial cities, unemployment and economic hardships in regional areas by resettling people into the new colony of South Australia [12, 13]. The South Australian Act (1834) allowed the sale of land in the proposed settlement to individuals who would establish primary industries such as farming, mining and manufacturing [13-15]. Thus, emigration to South Australia was extensively advertised in Britain [16]. The vast size and natural environment of South Australia for farming, its Mediterranean climate and rich minerals could have attracted people to migrate to the new colony who wished to create and develop their own opportunies, rather than going to an established colony such as in North America [16]. The development of the new Australian colony required builders, mechanics, agricultural labourers, and miners [17]. A high number of migrants to South Australia came from the counties of Cornwall, Devon, Dorset and Somerset, followed by Lancashire, Middlesex, Staffordshire and Warwickshire [16, 18, 19].

Migrants who could not afford the cost of the voyage to South Australia could apply for an “assisted passage”. Migration via this route was regulated by The Colonial Land and Emigration Commission (CLEC), whose agents selected equal numbers of male and female healthy young people of “good character” [16, 17, 20]. The policy was designed to maintain a regular supply of skilled workers to landowners, new industries and to advance the development of new settlements [16, 17, 20]. The assisted passage to the new colony was funded by the British Government and The South Australian Company. Additional financial help, for individuals with an assisted passage, could be applied for from local charitable organisations. This money covered the cost of transport to the port of departure, the compulsory deposit required for bedding, utensils and a set of “all weather” clothing for the long voyage [16, 20].

In total, 186,054 indivduals migrated to South Australia between 1836 and 1900 from the United Kingdom. The government “assisted passage” was received by 123,039 (66%) migrants [17]. The above mentioned selection criteria were not applied to individuals who paid the full costs of their passage and had adequate funds to support themselves in the colony.

The health of migrants on arrival in South Australia was of a high priority for the British Government. Therefore, the Colonial Land and Emigration Commission monitored conditions on board ships. Each ship had a Surgeon Superintendent, who was accountable for the health and wellbeing of all passengers [16, 20, 21].

Political disagreements within the new South Australian Government delayed the surveying of land for migrant settlements and the development of supporting infrastructure [12]. This affected the initial establishment of farms, food production, industrial enterprises, and permanent housing for migrants. These issues also delayed the development of the economy, caused unemployment and led to the removal of the first Governor, Captain John Hindmarsh. The second Governor, Lieutenant Colonel George Gawler, was appointed by the British Government in 1838 [19, 22]. Governor Gawler had a proactive approach to leadership and commissioned multiple infrastructure projects to rapidly develop the colony [22, 23]. Payment of the expenditure associated with these developments was rejected by the British Government, who were unwilling to cover the high costs, and recalled Governor Gawler to London in 1841 [19, 24].

The third South Australian Governor, Captain George Grey, was appointed the same year, and the colony faced its first economic depression [19, 25]. The lack of funds to continue public works meant Governor Grey faced retrenchment between 1841 and 1845. He redirected the majority of the unemployed workforce to agricultural industries [12, 19]. The eventual outcome was an increase of agricultural goods, particularly wheat and animal products and the establishment of export industries to other Australian colonies and Britain [12, 15].

Climatic conditions of South Australia, high summer temperatures and limited rainfall, often resulted in periods of drought and poor harvests. These conditions could have contributed to the poor economic growth and the unemployment experienced during the establishment of the colony. Throughout this period, the life of many settlers was harsh, particularly those who were unemployed and had to depend on charitable organisations and the government for their survival [23]. Government of South Australia State Records [26], indicate that 446 sick and destitute people received help in the form of food rations from the Emigration Department in 1839 - 1840. This number increased to 904 persons for the period of 1840 - 1841 [23]. In 1843, an act was passed relating to the care of the needy and this was followed by the establishment of the Destitute Board in 1849. This board offered support to the elderly, chronically infirm, and some widows, and initiated the construction of a ‘Destitute Asylum’ in the city of Adelaide during the 1850s [23, 27, 28].

The Destitute Asylum was modelled on the British workhouse system [27], with strict regulations, such as complusory wearing of uniforms for “inmates”, and severe penalties if regulations were not followed [23, 29]. This “indoor” relief at the Destitute Asylum was the last resort for individuals who had to prove that they had no relatives able to support them [29]. Expectant mothers with no other support were also admitted to the asylum for up to six months and received “Lying-In-Home” relief [23, 29]. Furthermore, “deserted” women with children, who had no means of support were allowed admision to the asylum [29].

Individuals, who still had their own accommodation but no other support, were given weekly food rations and fire wood as “outdoor” relief from the Ayslum [29]. People who lived in rural areas either had to travel long distances into the city of Adelaide to receive “outdoor” relief or cope as best they could within their own community.

Other burdens that early migrant settlers had to face were the spread of infectious diseases such as diphtheria, typhoid, typhus fever, and tuberculosis [30-34]. In addition, individuals who could not afford the cost of local medical services would have been badly affected, as the only public hospital was located at the city of Adelaide [35, 36]. People, who lived in rural areas may also have had limited access to health services [36]. Chronic conditions such as metabolic deficiencies, together with some of the above-mentioned diseases, can cause changes in the bone morphology. A number of skeletal abnormalities, for example, abnormal porosity in the cortex of bones, bending of long bones, enlargement and flaring of costochondral junctions of ribs, and porous lesions of the bones of the orbital roof, have previously been interpreted as signs of metabolic deficiencies, [37-41]. However, these skeletal lesions can be produced by a range of aetiologies [42-44].

St Marys-on-the-Sturt was an early rural migrant settlement, which was established from the late 1830’s [45]. It was a small village located eight kilometres south of the city of Adelaide (Fig.1A to C). The majority of the settlers were of British origin, who wished to establish an Anglican Church (St Mary’s) in the village. They also allocated the land surrounding the church for a cemetery (Fig.1D), with the first burial in 1847 [46]. The cemetery had a section referred to as the “free ground”, where individuals or their families, who had no funds to support the cost of a burial were interred. The burials in the “free ground” were unmarked and the costs were paid for by the Government of South Australia [23, 47, 48]. This section of the cemetery was located at the rear of the church building (Fig. 1D) and was in use from 1847 to 1927. [45]

**Fig 1.**
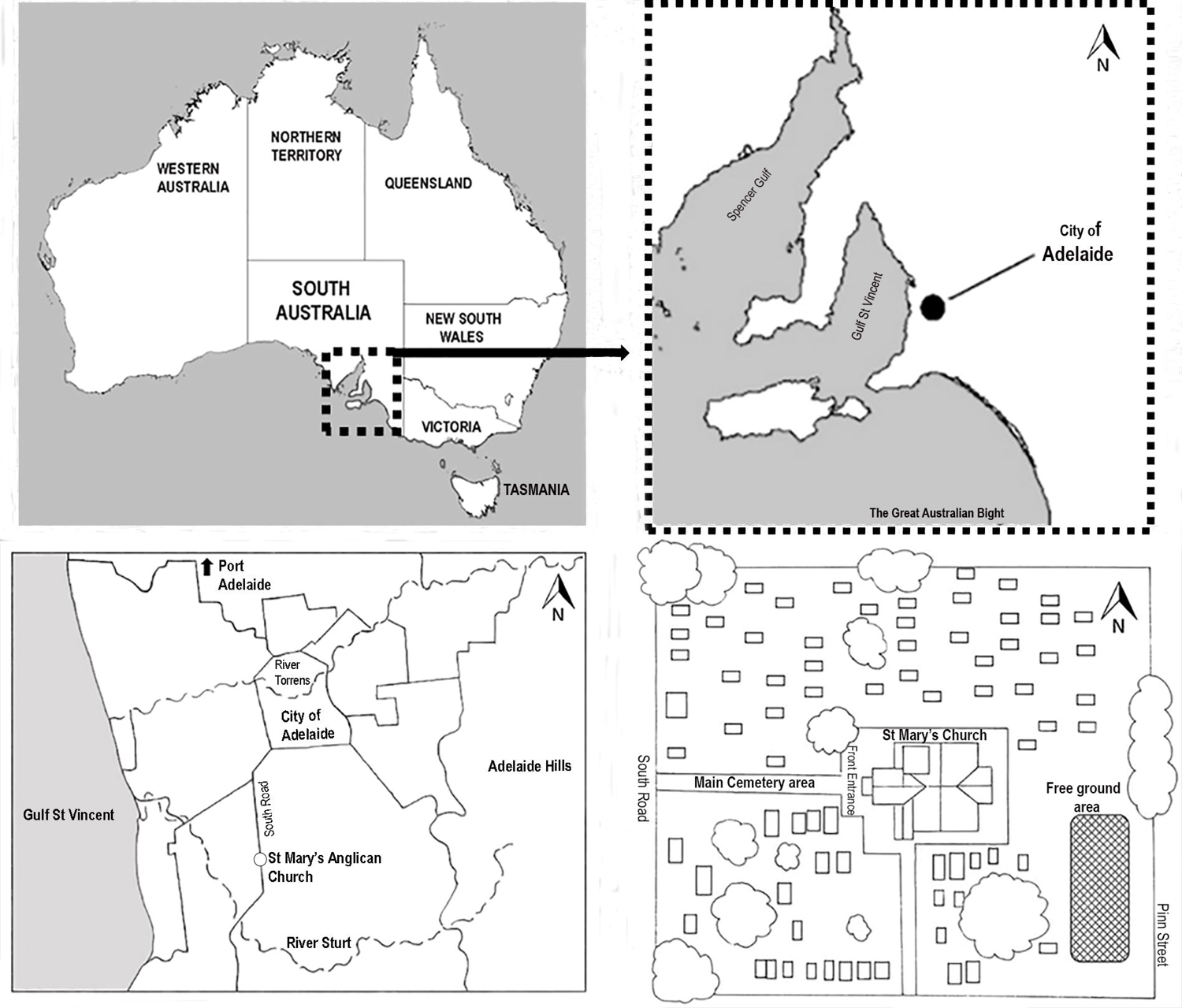
Location maps. **A:** Location of South Australia in relation to other Australian States and Territories. **B**: Location of the city of Adelaide, capital of South Australia. **C**: Location of St Mary’s Anglican Church Cemetery in relation to the city of Adelaide. **D**: *St Mary’s Church and associated cemetery. The hatched rectangle shows the “free ground” area which is located at the rear of the church building*.

The focus of this study was to investigate abnormalities seen in the skeletal remains excavated from the “free ground” area of St Mary’s Anglican Church Cemetery and understand the lifeways of these people. This study cannot ascertain the socioeconomic status of each individual buried in the “free ground” of St Mary’s Cemetery from the observed skeletal manifestations. However, the location of their burial suggests they had faced economic hardship around the time of death. Parish records of the St Mary’s Church Cemetery, list occupations of individuals who were buried in the “free ground” area, as, among others, cabinet maker, blacksmith, schoolteacher, and accountant. Some of these people had skilled trades and were considered to be more than just labourers. The probability is that this was a mixed group, mainly people who were from a working class.

## Aims and Objectives

The aims of this study were to

1. Investigate manifestations of disease in the skeletal remains of a group of settlers buried in the “free ground” area of St Mary’s Anglican Church Cemetery, South Australia, from 1847 to 1927.
2. Compare the findings of the St Mary’s samples with those published for skeletal samples from two 19^th^ century British communities.
3. Compare the number of individuals buried during the study period (1847 to 1927) in other sections of St Mary’s Cemetery to the number of people and the percentage of burials at the “free ground” area to understand the economic context of life in the new colony.

## Material and Methods

Skeletal remains of 65 individuals (20 adults and 45 sub-adults), excavated from the “free ground” section of St Mary’s Anglican Church Cemetery, South Road, Adelaide, South Australia (Site code: SMB - St Mary’s Burial) in 2000, were available for examination. No other sections of the cemetery were excavated. These individuals were buried in the “free ground” between 1847 and 1927 in unmarked graves, which prevented identification of individuals. Sex and age at death were estimated immediately post excavation using the standard physical anthropological methods [47]. The skeletal remains are part of an archaeological collection; therefore, no destructive analysis was permitted.

### Scoring of skeletal material

Each skeleton was arranged in the anatomical position, and an inventory of the bones was prepared as described by Buikstra et al.[49] and Mitchell and Brickley [50]. Skeletal manifestations related to metabolic deficiencies were recorded as described by Brickley[51], Brickley and Ives [37], Heron [52], Mays et al., [39], Ortner et al., [41, 53], and Ortner & Ericksen,[40] (Table 1 and Table 2). Results that are presented in Table 1 are shown as the number of individuals with the observed manifestation (-n-) over the total number (-N-) of individuals with the bone available for observation (n/N).

**Table 1.** Prevalence of observed skeletal manifestations associated with health deficiencies observed in different bones of the skeletons of individuals from St Mary’s sample.

| Age (years) | 0-1 | 1-4 | 5-9 | 10-14 | 15-19 | 20-29 | 30-39 | 40-49 | 50-59 | 60+ | Total | Total Prev. % |
| --- | --- | --- | --- | --- | --- | --- | --- | --- | --- | --- | --- | --- |
| **Abnormal porosity in the cortex of bones listed below:** |  |  |  |  |  |  |  |  |  |  |  |  |
| **Maxillae:**<br>posterior surface | 1/5 | 2/11 | 0/2 | 0/2 | 0/1 | 0/1 | 0/2 | 0/3 | 0/2 | 0/1 | 3/30 | 10 |
| Palatine processes | 1/4 | 0/13 | 0/2 | 0/3 | 0/1 | 0/1 | 0/3 | 1/5 | 1/6 | 0/1 | 3/39 | 8 |
| Alveolar process | 2/3 | 4/13 | 1/2 | 1/2 | 0/1 | 0/1 | 1/3 | 0/5 | 0/6 | 0/1 | 9/37 | 24 |
| **Mandible:**<br>Coronoid process | 1/9 | 0/13 | 0/2 | 0/3 | 0/1 | 0/1 | 0/3 | 1/5 | 0/6 | 0/1 | 2/44 | 5 |
| Alveolar process | 1/9 | 0/15 | 0/2 | 1/3 | 0/1 | 0/1 | 0/3 | 0/5 | 0/7 | 0/2 | 3/48 | 6 |
| Greater wing of the sphenoid bones | 0/3 | 1/6 | 1/3 | 1/2 | 0/1 | 0/1 | 0/3 | 0/5 | 0/6 | 0/2 | 3/32 | 9 |
| **Other skeletal changes to bones listed below:** |  |  |  |  |  |  |  |  |  |  |  |  |
| **Ribs:**<br>Enlargement costochondral junctions | 1/4 | 0/5 | 0/1 | 0/2 | 0/1 | 0/1 | 0/3 | 0/3 | 0/2 | 0/0 | 1/22 | 5 |
| **Long bones:**<br>Flaring-distal metaphysis | 0/13 | 1/14 | 0/3 | 0/1 | 0/1 | 0/1 | 0/3 | 0/5 | 0/7 | 0/1 | 1/49 | 2 |
| **Orbital roofs porous lesions (any Type)** | 0/7 | 2/14 | 1/3 | 3/3 | 1/1 | 0/1 | 0/3 | 0/5 | 2/8 | 0/2 | 9/46 | 20 |
| **Orbital roofs porous lesions (Types 3 or 4)** | 0/7 | 1/14 | 1/3 | 1/3 | 0/1 | 0/1 | 0/3 | 0/5 | 0/6 | 0/2 | 3/46 | 7 |

**Note.** Results presented as the number of individuals with the observed sign (-n-) over the total number (-N-) of individuals with the bone available for observation (n/N), with age groups of the individuals.

**Table 2.** St Mary’s sub-adult samples, with their age range, showing two or more macroscopic pathological manifestation of a health deficiency.

| Burial Code | SMB 27B | SMB 28 | SMB 51 | SMB 56 | SMB 58 | SMB 70 |
| --- | --- | --- | --- | --- | --- | --- |
| Age range at death (years) | 0 to 5 | 10 to 15 | 10 to 15 | 0 to 5 | 0 to 5 | 7 to 12 |
| Greater wing of the sphenoid bones † | -- | P | -- | -- | A | P |
| Zygomatic bones: internal surface † | -- | -- | -- | -- | P | -- |
| Maxillae: posterior surface † | A | A | -- | P | P | A |
| Maxillae: palatine process † | A | A | A | P | A | A |
| Maxillae: alveolar process † | P | P | -- | P | P | P |
| Mandible: coronoid process medial surface † | A | A | A | P | A | -- |
| Mandible: alveolar process † | A | A | P | P | P | -- |
| Orbital roofs: †<br>Type: (number given) | P 1 | P 4 | P 1 | -- | A | A |
| Scapulae: †<br>supraspinous or infraspinous areas | -- | A | A | P | A | P |
| Pelvic bones † | A | A | A | P | A | P |
| Ribs: enlargement of costochondral junctions | -- | A | A | P | A | -- |
| Long bones: Flaring of distal metaphyses | -- | A | -- | A | A | A |

**Key:** † = Areas of abnormal porosity (lesions) in the bone cortices observed
**P** = condition present
**A** = condition absent
**--** = condition unobservable due to skeletal part missing

Porous lesions on the bones of the orbital roof were quantitated using the scoring system described by Stuart-Macadam [54], i.e., Type 1 - “capillary-like impression on bone”, Type 2 - “scattered fine foramina”, Type 3 to 5 - ranged from “large and small isolated foramina to outgrowths from trabecular bone that extended to the surface of the outer table.

Investigation of the internal structure of tooth samples for interglobular dentine (IGD) was part of the study. The traditional methods used for such investigations were histological techniques and these require sectioning of the tooth sample. Consequently, a non-destructive method introduced to detect IGD, using X-Ray Computed Tomography (micro-CT) was used. Cost of this method allowed only a selection of tooth samples from St Mary’s skeletal collection to be investigated.

One tooth each from 19 individuals was selected. The individuals were selected from a broad age range (∼2 years to 60+ years of age). Collected teeth included two permanent incisors, three permanent canines, three permanent premolars, nine permanent first molars, and two primary molars, as the same tooth type was not available from each individual. Each tooth was scanned using the Bruker SkyScan 1276 Micro-CT scanner at Adelaide Microscopy, The University of Adelaide [55]. The scanner was set at source voltage: 100 kV, source current 200 µA, camera binning: 4032 x 2688, filter: aluminium and copper, pixel size: 9.0 µm. Tooth sample from SMB 63, was additionally scanned using the pixel size was 5.21 µm.

Micro-CT scan datasets were reconstructed into a visual image using NRecon, a volumetric reconstruction software. These reconstructed scan data sets were viewed as either two-dimensional (2D) or three-dimensional (3D) images using Dataviewer, a ‘volume rendering’ software and Avizo 9 software [56]. The 2D and 3D images were analysed to identify mineralisation defects in the teeth. Dentine defects seen on the micro-CT scans were scored, as described by Colombo et al. [57] and Veselka et al. [58].

### St Mary’s Parish Records and Cemetery Survey

Parish records from St Mary’s Anglican Church, South Australia, in relation to potential burials in the “free ground” area of the cemetery from 1847 to 1927 were used. These documents recorded the burial location of some individuals, but not all [47]. The parish records also provided an approximate date of burials and the occupation for some individuals. Calculations for Figure 3 used the data from these records.

The date of death and approximate date of burial for individuals who paid for their burial in provenanced graves, i.e., in the “leased ground” of the main section of the cemetery with a memorial marker/gravestone (Fig. 1D), was recorded during a survey of the St Mary’s Cemetery.

### Comparison of observed metabolic deficiencies from St Mary’s samples with those of British samples

Findings of the St Mary’s samples were compared with those published for two 19^th^ century to early 20^th^ century British skeletal samples. One sample was from St Martin’s-in-the-Bullring Church, Birmingham, England (N= 406) [37, 39, 59], where the majority of burials were between 1810 and 1864, with declining numbers of individuals interred until 1915 [59].

St Martin’s was located in an industrial city where many individuals from the working classes might have been buried, this sample was considered appropriate as many individuals interred in the “free ground” of St Mary’s, or their families if they were infants, were likely from a similar socioeconomic working-class background at the time of death.

The published results for a second Bristish skeletal sample were from a section of St Peter’s Collegiate Church, Wolverhampton, England, referred to as the “overflow burial ground” (1819 to approximately 1900) [60, 61]. Wolverhampton was originally a market town with a mix of agriculture and small industries. The industrial development of local mining activities, during the 19^th^ century, contributed to the increase in population size [60]. Published findings from St Peter’s skeletal samples were considered an appropriate comparison sample to the St Mary’s sample as some individuals in St Peter’s sample could have been miners, and it is known that many migrants to the new colony were from mining communities [16, 17, 62, 63].

## Results

### Demography

Estimated age at death range and estimation of sex for the skeletal samples from St Mary’s ‘free ground” are shown in Figure 2. Among the 45 sub-adults in this sample, 36 were under two years of age.

**Fig 2.**
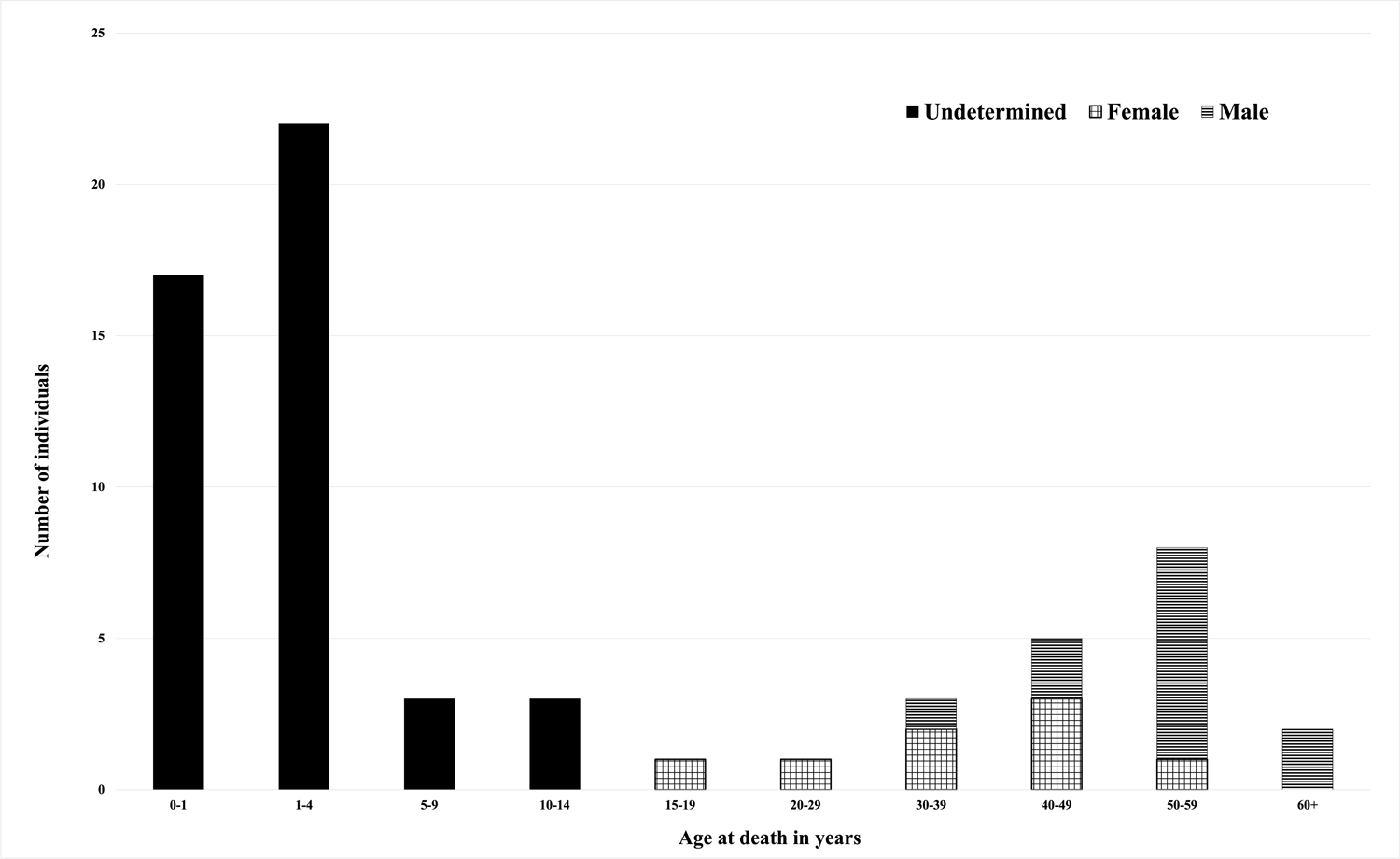
Demography. Distribution of the excavated St Mary’s “free ground” skeletal samples by age at death and sex [47].

### Parish Records and Cemetery Survey Data

In the main cemetery area, surrounding St Mary’s Church, a total of 227 people were buried in “provenanced” graves between 1847 and 1927. The comparison of the number of individuals recorded in parish records as *possibly* buried in the “free ground” area of St Mary’s Cemetery (n=195), with the number of provenanced burials (n=227), by the decade of burial are presented in Figure 3.

**Fig 3.**
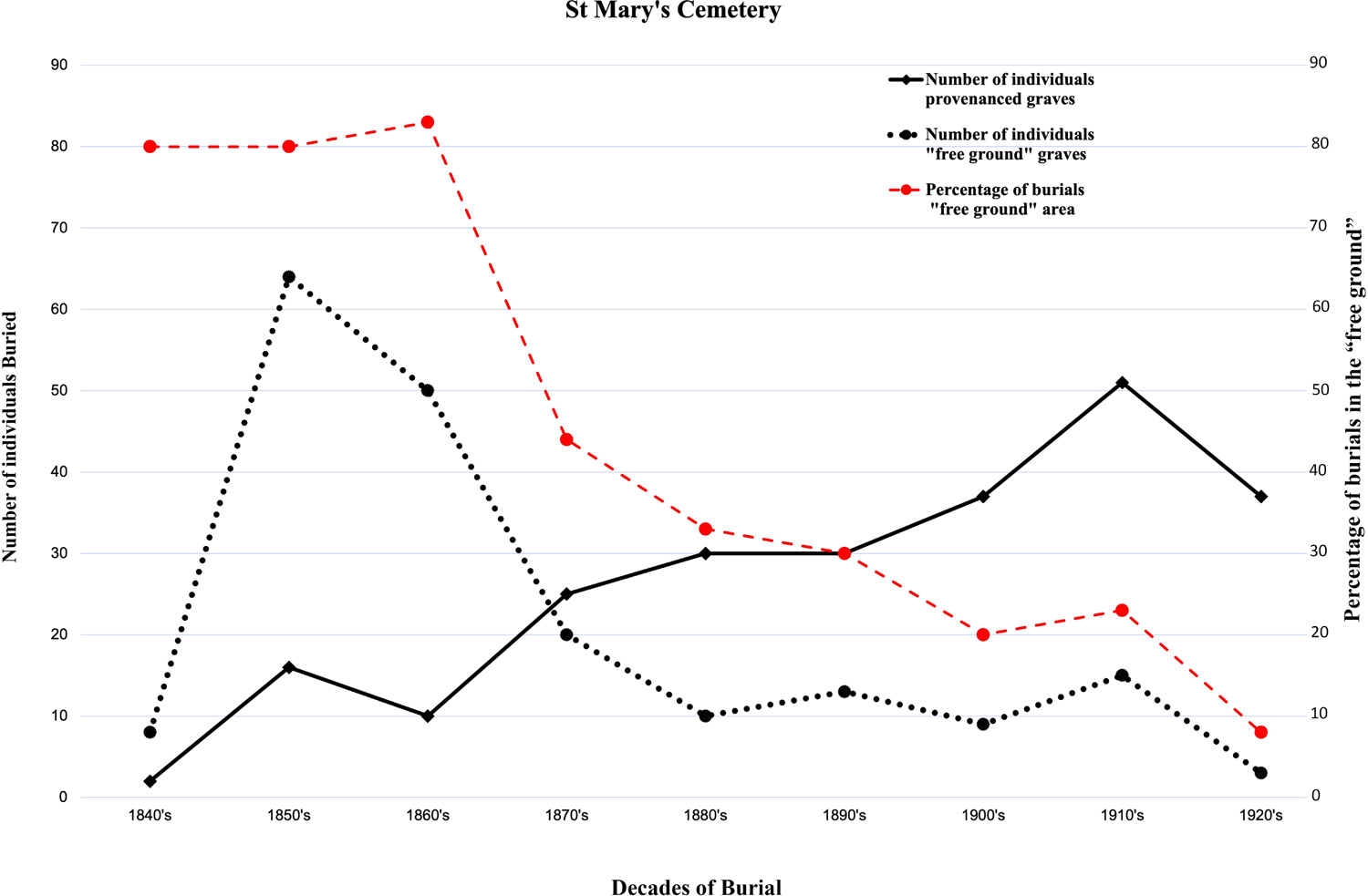
St Mary’s Parish Records. A Comparison of the number of people possibly buried in the “free ground” area (n=195) with those buried in provenanced graves (n= 227) of St Mary’s cemetery, according to parish records [47], by decades of burial. Red **dashed line** indicates the percentage of burials in the “free ground” area of the cemetery.

### Summary of observed skeletal manifestations

Pathological manifestations relating to health deficiencies, possibly metabolic, were identified on the skeletal remains of some individuals buried at St Mary’s “free ground” area. They are presented below:

### Abnormal porosity in the bone cortex

Nine adults and 12 sub-adults were observed with at least one area of abnormal porosity in the cortical parts of following bones: maxillae – the posterior surface, alveolar process, and palatine processes (Fig. 4); medial surfaces of the coronoid process of the mandible; and the greater wing of the sphenoid. One infant, SMB 56, (approximately 6-9 months of age), in addition to the above bones, showed areas of abnormal porosity in cortices of the lateral and basilar portion of the occipital bone, scapulae, ribs, vertebral arches, ilia, and the extremities of long bones. The prevalences of abnormal porosity are presented in Table 1.

**Fig 4.**
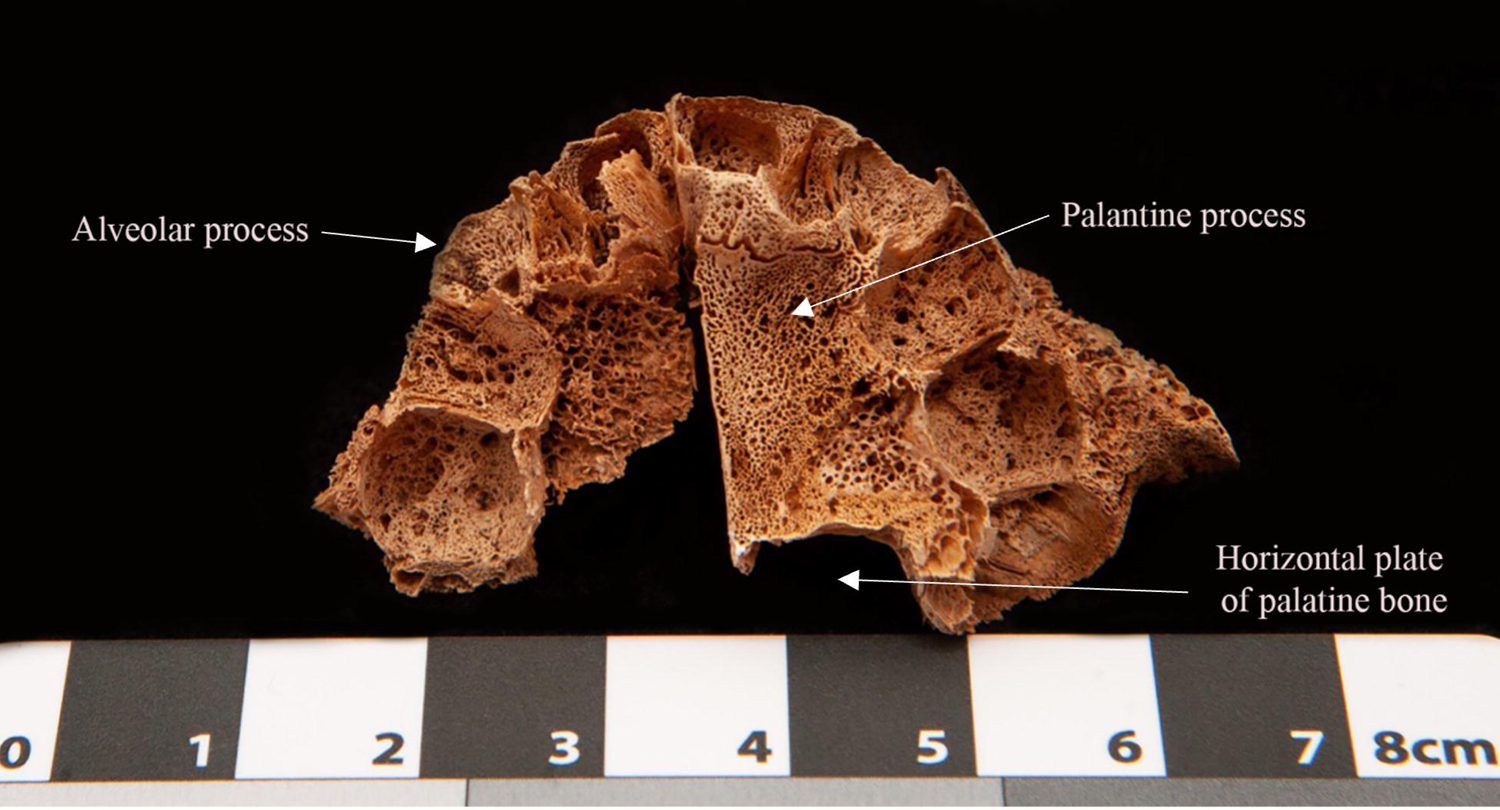
SMB 56. Palate, inferior view, showing areas of abnormal porosity that extended throughout the alveolar process of the maxillae.

### Enlargement and flaring of the costochondral junctions of ribs

One infant, SMB 56, had enlargement and flaring of the costochondral junctions of ribs (Table 1).

### Enlargement and flaring of the metaphyses

One sub-adult, SMB 8, showed flaring and enlargement of the distal metaphyses of the femora (Table 1).

### Porous lesions on the bones of the orbital roof

Three sub-adults, SMB 4A, SMB 19 and SMB 28, age range from 4 years to 13 years, showed porous lesions on the bones of the orbital roof of Types 3 and 4 [64], which are often referred to as ‘cribra orbitalia’. The lesions of SMB 28 were composed of small and large pores on the right and left orbital roof respectively (Fig. 5). The pores on the right orbital roof seemed to penetrate the cortical bone, while those on the left appeared to be exposed trabecular bone (Fig. 5). This sub-adult also displayed areas of abnormal porosity in the cortex of the greater wing of the sphenoid bones bilaterally (Table 1). The prevalence rate of porous lesions seen on orbital roof bones in St Mary’s sample are presented in Table 1.

**Fig 5.**
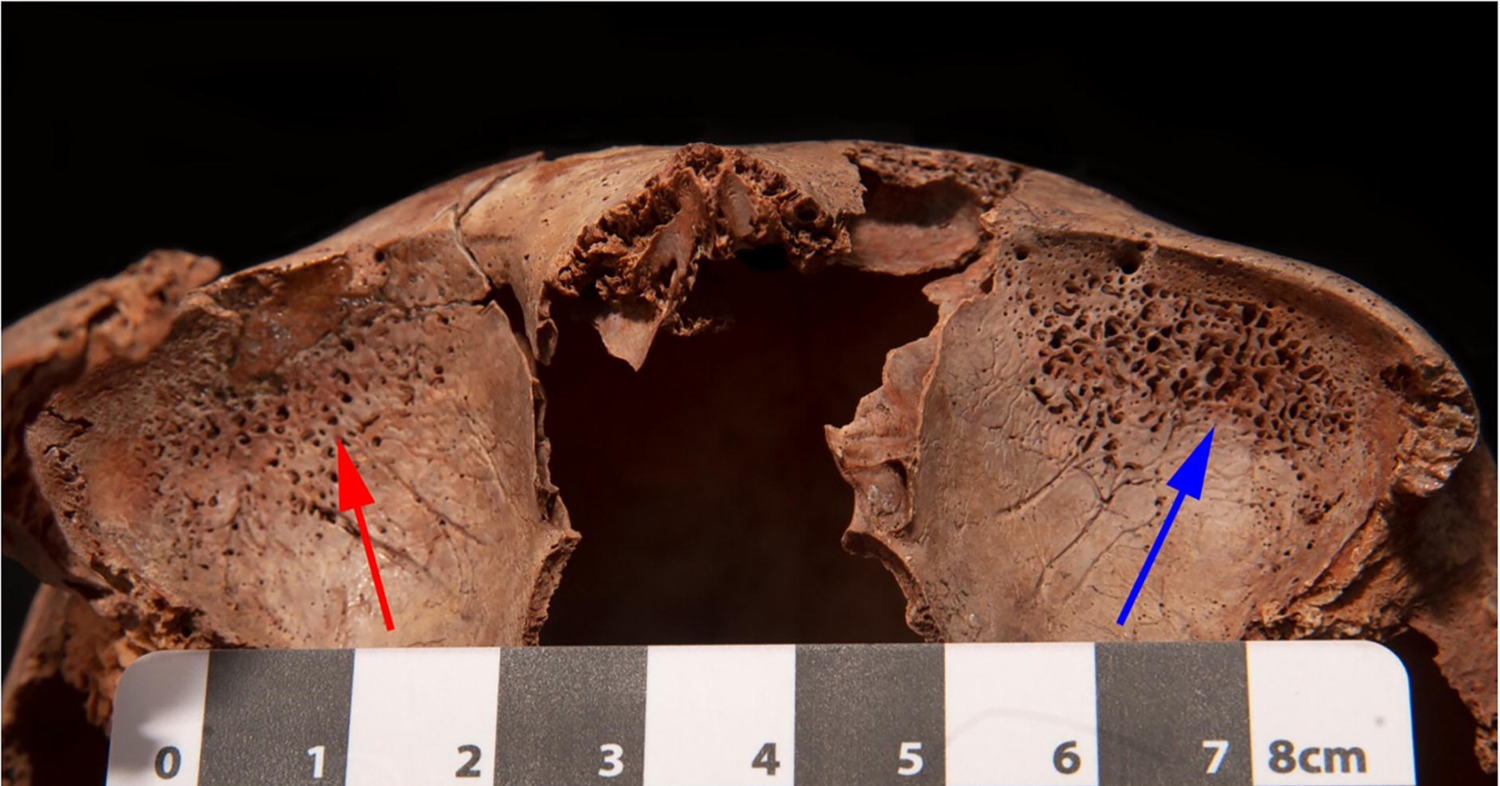
**SMB 28**. Lesions on the orbital roof bones. Red arrow indicates small pores in the right cortical bone. Blue arrow indicates possible exposed trabecular bone on the left.

### Dentine - Interglobular dentine (IGD)

Lesions in the internal structure of the tooth seen in micro-CT scans were areas of unmineralised collagen matrix in the dentine. Such lesions have been described as interglobular dentine [65, 66]. Tooth samples from three individuals, SMB 6 (∼ 40 to 45 years of age), SMB 63 (∼55 to 60 + years of age), and SMB 70 (∼ 7 to 12 years of age) (Fig. 6) were seen with areas of IGD. Individual SMB 70 had suffered from congenital syphilis and had been treated with mercury [67]. Therefore, this sub-adult was excluded from this part of the study as the toxic treatment could have resulted in IGD. The tooth sample from SMB 63 (a permanent lower lateral incisor) was selected for further investigation using a higher resolution of the micro-CT scanner (pixel size: 5.21 µm). This revealed areas of IGD in three separate incremental linear arrangements, one of the areas of IGD was observed opposite an external enamel hypoplastic defect (Fig. 6). The crown of this tooth type commences mineralisation at approximately 30 weeks of intrauterine life (±1 month) and completes at approximately 3.5 years (±1 year).

**Fig 6.**
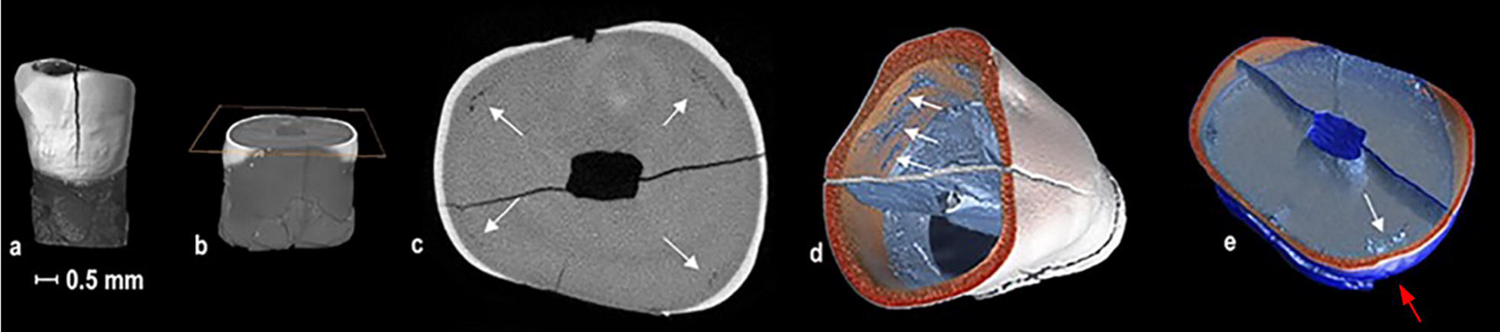
Micro-CT images. SMB 63. (a) Lower lateral permanent incisor with a fracture in crown. (b) Transverse slice: at the level of IGD in the crown. (c) Transverse slice: arrows show four areas of IGD. (d) Transverse slice: arrows show three areas of IGD, image was angled to show location of IGD in three concentric layers. (e) White arrow shows IGD (internal) opposite an enamel hypoplastic defect red arrow (external).

Sub-adults from the St Mary’s sample who showed two or more macroscopic pathological manifestations of a health deficiency are listed in Table 2.

Findings of St Mary’s samples and demographic profiles were compared with published results for 19^th^ century skeletal samples buried at St Martin’s-in-the-Bullring Church, Birmingham, England, and St Peter’s Collegiate Church, (overflow burial ground), Wolverhampton, England. A comparison of the number and percentage of adults and sub-adults from each sample is presented in Table 3.

**Table 3.**
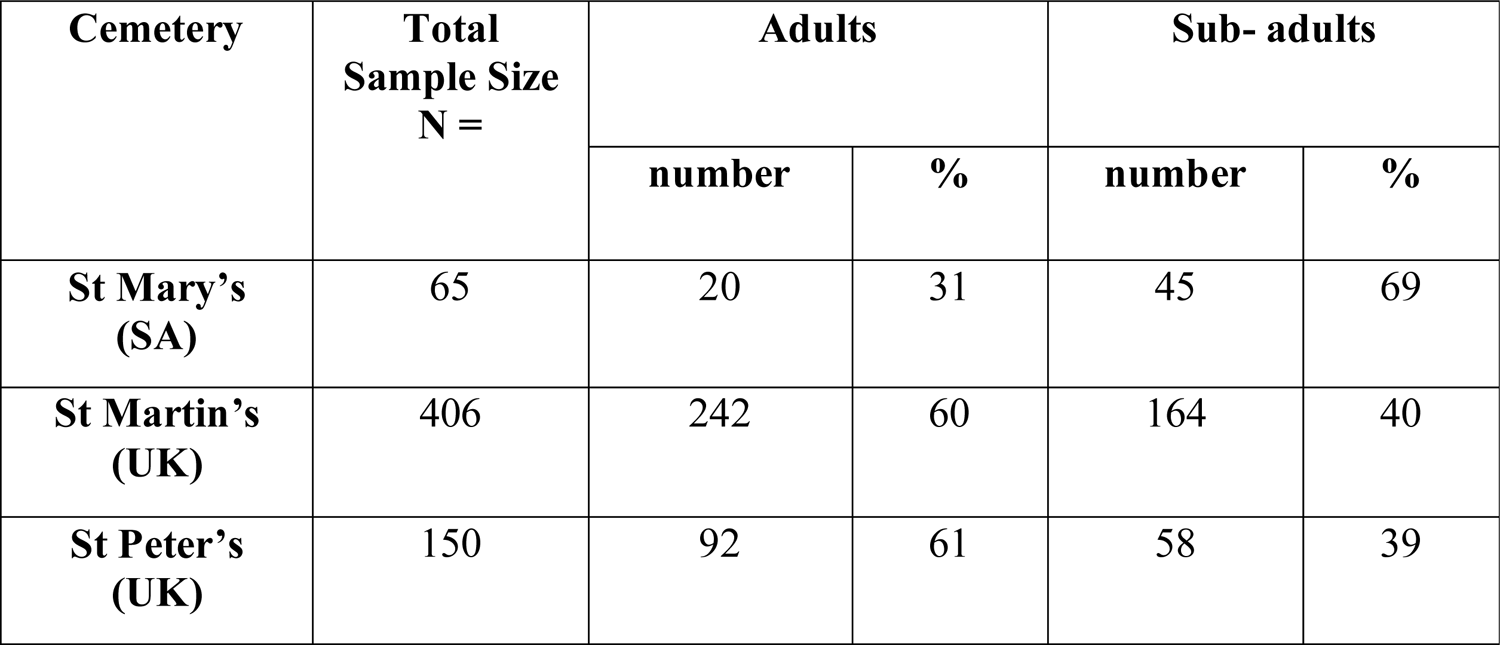
Demographic profiles of St Mary’s, St Martins and St Peter’s cemeteries

| Cemetery | Total<br>Sample Size<br>N = | Adults |  | Sub- adults |  |
| --- | --- | --- | --- | --- | --- |
|  |  | number | % | number | % |
| St Mary's<br>(SA) | 65 | 20 | 31 | 45 | 69 |
| St Martin's<br>(UK) | 406 | 242 | 60 | 164 | 40 |
| St Peter's<br>(UK) | 150 | 92 | 61 | 58 | 39 |

A comparison of the macroscopic observed skeletal manifestations among the sub-adult samples from the three cemeteries is presented in Figure 7.

**Fig 7.**
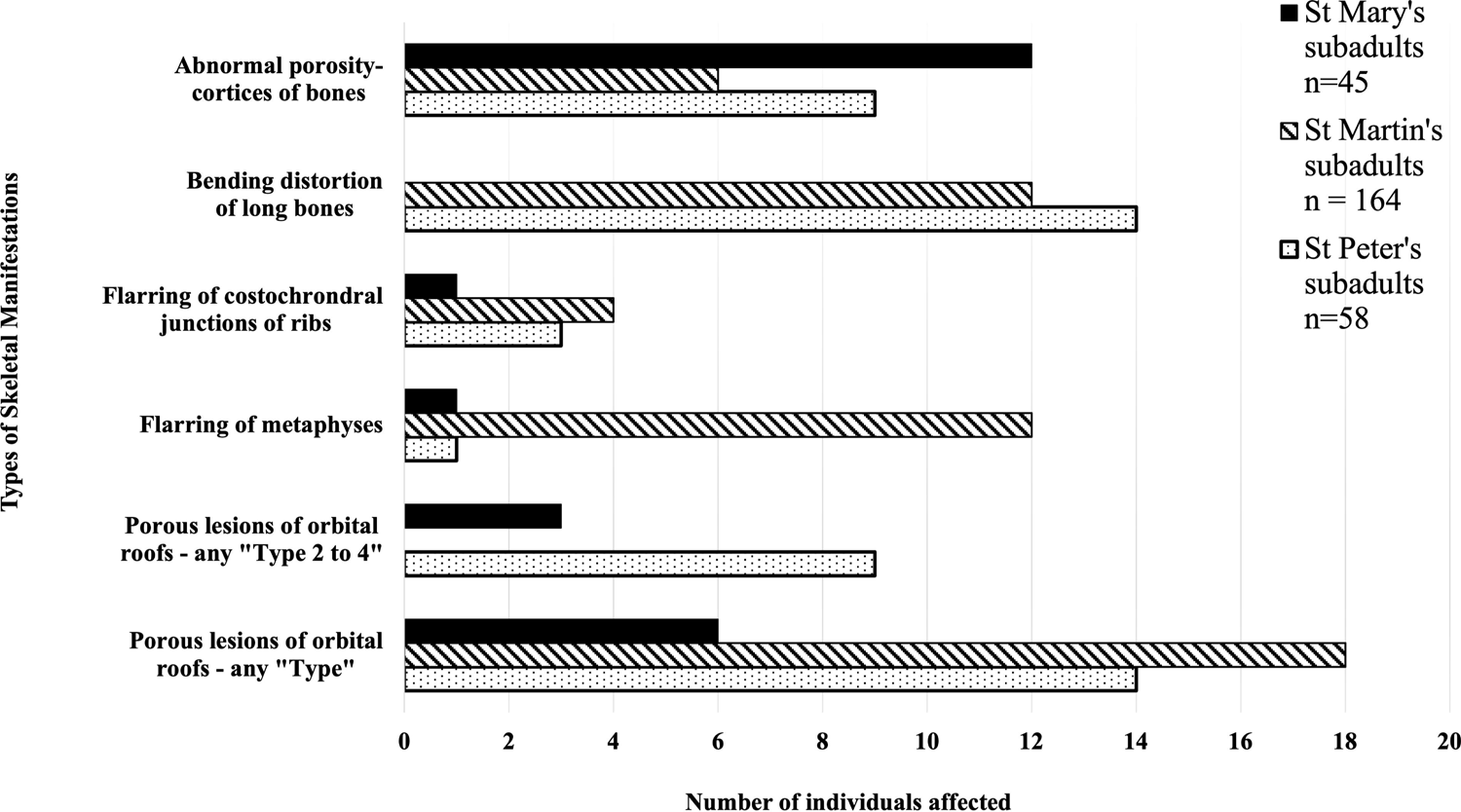
**Comparison of the three cemeteries**. Skeletal manifestations observed in the sub-adults samples from St Mary’s, St Martin’s and St Peter’s Cemeteries and the number of individuals affected.

### Abnormal porosity in the bone cortex

Sub-adults from St Martin’s and St Peter’s Cemeteries were observed with areas of abnormal porosity in the cortices of the following bones: infra orbital area, maxillae, hard palate, coronoid processes of the mandible, orbital, parietal and occipital bones, and scapulae [37, 59, 61]. St Mary’s had more sub-adults with this manifestation (abnormal porosity in the bone cortex) compared to St Martin’s and St Peter’s samples (Fig. 7). Nine adults from the St Mary’s sample were also seen with one area of abnormal porosity in the cortices of bones. However, this skeletal manifestation was not observed in any adults from St Martin’s or St Peter’s samples [37, 59, 61].

### Bending /bowing distortion of long bones

This skeletal abnormality was seen only among sub-adults of St Martins and St Peter’s cemeteries (Fig. 7) [39, 59, 61]. Furthermore, no adults from either of the three cemeteries were seen with this skeletal abnormality.

### Enlargement and flaring of metaphyses

St Martin’s had a greater number of sub-adults with flaring of metaphyses than St Mary’s and St Peter’s sub-adult samples (Fig. 7) [39, 59, 61]. No adults from the three cemeteries were seen with this skeletal manifestation.

### Porous lesions on the bones of the orbital roof

The scores for this specific skeletal manifestation from St Martin’s samples cannot be compared between St Mary’s and St Peter’s samples as they were not available separately for the Types 3 and 4. However, published results for this cemetery do state that 18 sub-adults and 17 adults were observed with “varying degrees” of porous lesions on the bones of the orbital roof (Fig. 7). [59].

## Discussion

The “free ground” area of St Mary’s Anglican Church Cemetery was allocated for the interment of people from the community of St Marys-on-the-Sturt, South Australia, who did not have the funds to cover the cost of a burial. According to the parish records from St Mary’s Church, for the decades: 1840s, 1850s, and the 1860s; 80%, 80%, and 83% respectively, of the total number of people buried at the St Mary’s Cemetery were interred at the “free ground” area (Fig. 3). This indicates that for approximately 30 years, over 80% of the individuals who were buried at the cemetery or their families, could not pay for the burials. This was the period of establishment of the new colony, during which time, the colony experienced economic recession and had a high unemployment rate [12, 19].

Therefore, a good percentage of the early settlers at St Marys-on-the-Sturt region may not have had regular employment to earn an adequate amount of money to support themselves and save for the latter part of their life. They may have had to depend on charitable organisations or the Government for their survival. This early economic hardship in the colony is reflected in the need for relief from the Destitute Asylum for many living in or near the city of Adelaide. Introduction of an old age pension, after the turn of the 20^th^ century, reduced the need for the Destitute Asylum [29].

Thirty six out of the 45 subadults (80%) buried at the “free ground” area were less than two years of age (Fig. 2). St Mary’s Church parish records list some infants as “unbaptised - no service” [47]. It is possible that some of these infants were stillborn or died a short time after birth. Such infants could have contributed to the high percentage of sub-adults in the St Mary’s sample. There were no official regulations regarding the location of burial of a still born infant in the early colony or the requirement to register the birth of a still born child until 1936 [48, 68]. Burial practices of perinates, excavated at the 19^th^ century Parramatta Convict Hospital in NSW, Australia, showed they were afforded very little respect or care [69].

The percentage of burials at the St Mary’s “free ground” area began to reduce from the 4^th^ decade after the establishment of cemetery and reached 8% of the total burials in the decade of the 1920’s (Fig. 3). This demonstrates that there was a gradual improvement of the economic status of some individuals who lived at St Marys-on-the-Sturt region. This local improvement could have followed the national trend of economic development of the colony of South Australia. This also shows that the majority of the skeletons studied here come from the earlier period in the life of the colony and practically characterise its health status during the 19th century, rather than reaching into the 20^th^ century.

Bones of the skeleton are a dynamic tissue and undergo remodelling during life, possibly according to the variations in forces acting on them during life. Therefore, disease manifestations seen in skeletal remains, particularly in bone cortices, occurred during the last remodelling before death [70, 71].

Abnormal porous lesions on bone cortices result from defective calcification of the bone matrix [40, 41, 51, 72]. Vitamin C and vitamin D deficiencies affect collagen synthesis [73, 74], and affect mineralisation [65, 75, 76], respectively. Abnormal porosity of the bone cortices seen in archaeological skeletal samples could be produced by both processes. In addition, porous lesions seen on the bones of the orbit roof could be caused by an anaemia. Anaemia has often been associated with a deficiency of iron. [54, 77, 78].When haemoglobin becomes inadequate, due to the lack of iron in the body, the red marrow compensates by the overproduction of red blood cells and thus produces a proliferation of red bone marrow. This can result in an expansion of the trabecular bone, amongst other pathological processes, between the tables of the skull [78]. In extreme conditions, the surface bone cortex of the orbital roof may thin out and expose the underlying trabecular bone, which could appear as porous lesions.

Abnormal porous lesions that were seen in the cortical bone of 9 adults and 12 sub-adults of St Mary’s skeletal collection could have resulted from either vitamin C or D deficiencies (Table 1). Presence of abnormal porosities in the bone cortices of the greater wing of the sphenoid, bilaterally, has been considered in the past as pathognomonic of vitamin C deficiency [40, 41]. Three of St Mary’s sub-adults would fit into this category (Table 1).

However, Brickley [38] has suggested that abnormal porous lesions should be present on more than one bone/location of the skeleton to be considered as a diagnosis of vitamin C deficiency. Accordingly, six of St Mary’s sub-adults including two of the three sub-adults mentioned above with lesions on the sphenoid bones, would fit into the “multiple” lesions category (Table 2). Furthermore, one sub-adult in this category had flaring of the costochondral junctions of the ribs. This skeletal manifestation could be attributed to either vitamin C or vitamin D deficiencies (Brickley 2008). The remaining nine adults and six sub-adults only had one area of abnormal porosity on one location of the skeleton. Therefore, could not be considered as individuals with a deficiency of vitamin C. The areas of abnormal porosity seen on these individuals could represent a manifestations of vitamin D deficiency, [38, 39]. However, none of them had any other of the common signs of vitamin D deficiency, such as bowing and bending distortions of the long bones [38, 39, 79]. One sub-adult did have flaring of the distal metaphyses of the femora but without any other pathological manifestation observed elsewhere in the skeleton, this sub-adult could not be included in either of the above vitamin C or D categories (Table 1).

Three of St Mary’s sub-adults had porous lesions on the bones of the orbital roof of Types 3 and 4 [54]. These “types” of lesion have been considered as an indicator of anaemia. This could result from a dietary deficiency of iron, and/or vitamin B12, malabsorption of iron from the gut due to chronic diseases or gastrointestinal and blood parasites, chronic blood loss and/or genetic conditions or a combination of these conditions [42, 54, 78, 80-83]. One sub-adult, SMB 28, also had areas of abnormal porosity on the greater wings of the sphenoid bones, (bilaterally). Therefore, it is likely that this individual could have had a co-occurence of anaemia and vitamin C deficiency (Brickley, 2016; Fain, 2005; Ortner et al., 2001; Ortner & Ericksen, 1997). Vitamin C enchances the absorption of iron from the gut, so a deficiency of vitamin C in SMB 28 could have intensified any anaemia present.

Tooth development commences in utero and continues until approximately 21 to 23 years of age [84, 85]. In dentine, after the development of the tooth, the remodelling process is minimal compared to bone [65, 66, 86], therefore, any pathological changes (lesions) would remain for the rest of the life. The areas of IGD, seen in the two adults from St Mary’s sample, would have resulted from health insults that caused an interuption of the mineralisation process in dentine during the development of the tooth. It is unknown if these individuals spent their younger lives in the UK or South Australia, indicating that the IGD could have occurred before migration. The three areas of IGD seen in SMB 63 could have resulted from three separate episodes of health insults during their childhood. Furthermore, one of the health insults that caused an area of IGD may have also resulted in the enamel hypoplastic defect (Fig. 6.e) [87-89]. The formation of IGD has been widely related to vitamin D deficiency [57, 58, 90-93]. However, neither of the two St Mary’s adults showed any other skeletal pathological manifastations, including porous lesions or bending abnormalties of long bones.

### Comparison of St Mary’s “free ground” skeletal samples with those of St Martin’s and St Peter’s samples in Britain

The working-class background of some of the people buried at the “free ground” area of St Mary’s Cemetery could be similar to those published for St Martin’s and St Peter’s cemeteries in Britain [59, 61]. However, the demographic profile of the St Mary’s sample is somewhat different from those of the British samples, as the majority of St Mary’s sample is made up of sub-adults (Fig. 2). Most of these sub-adults were under two years of age and according to St Mary’s Church parish records, one of the common causes of death was gastrointestinal conditions [47]. Most of the sub-adult deaths could have resulted from dehydration resulting from diarrhoea or vomiting, as emergency access to medical help would have been difficult due to the location of the public hospital in the city of Adelaide, some 8 km away from St Marys-on-the-Sturt.

Areas of abnormal porosity in the cortices of bones, that could be considered as a deficiency of vitamin C, were seen in sub-adults from St Mary’s, St Martin’s and St Peter’s skeletal samples [37, 61]. The prevalence of such lesions was higher among the subadults from the St Mary’s sample (13 %) compared to that from St Martin’s (4 %) and St Peter’s samples (2 %). The lower prevalence of this manifestation (probable vitamin C deficiency) among the sub-adults of the British samples could be due to the availability of fresh fruits and vegetables in Birmingham and Wolverhampton. The initial difficulties and delays in establishing farms and the production of foods, during the first decades of the development of South Australia, indicates that foods particularly those rich in vitamin C, may not have been readily available. This was especially true if there was a scarcity of water [13, 94]. Some of the sub-adults affected by this deficiency, from all three of the cemeteries, could have been breastfed infants [37, 61]. Therefore, their deficienecy could have resulted from a dietary deficiency of the mother [51, 53, 95]. Infants could also have been affected by the feeding and weaning practices of the period [96-98]. Elevated nitrogen isotope values (+ 1.7‰) observed in skeletal remains of some infants from the St Mary’s sample [99], relative to those of adult females in the same sample, suggested that breastmilk was a principal source of diet for infants. However, these findings do not indicate whether the child received adequate amounts of milk during breast feeding.

The number of individuals with the manifestation of enlargement and flaring of the costochondral junctions of ribs and flaring of the metaphyses of long bones was higher among St Martin’s and St Peter’s sub-adult samples compared to those of St Mary’s samples (Fig. 7) [39, 61]. The manifestation of bending distortions of long bones was only seen among sub-adults of St Martin’s and St Peter’s samples, and not observed in any of sub-adult samples from St Mary’s (Fig. 7). Skeletal remains of adults from the three cemeteries were free of the above-mentioned manifestations. These skeletal abnormalities have been linked to a chronic deficiency of vitamin D [39, 61]. Among the St Mary’s subadults the lower incidence of the manifestations linked to vitamin D deficiency (Fig. 7) could be due to the abundance of sunlight (UV rays) in South Australia.

St Martin’s had the higher number of subadults with porous lesions in the bones of the orbital roof (Fig. 7) [59]. However, the scores for these porous lesions, as described by Stuart-Macadam [64], were not available (Brickley, personal communication, 2018). This lack of data made it difficult to compare St Martin’s to St Mary’s and St Peter’s samples in this aspect (Fig 7). Nevertheless, it is clear that many of St Martin’s sub-adults had experienced chronic systemic health insults before death [78].

A comparison between the South Australian and British samples assisted in the evaluation of the level of manifestations related to health deficiencies observed in the migrant settlers buried in the “free ground” area of St Mary’s Anglican Church Cemetery. It also contributes to a wider picture of life and death in the mid-19^th^ to early 20^th^ century community of St Marys-on-the-Sturt.

## Conclusion

Skeletal manifestations observed on individuals excavated from the “free ground” area of St Mary’s Anglican Church Cemetery, South Australia, show that St Mary’s sub-adults had a higher incidence of abnormal porosity on the bone cortex compared to the British samples. Furthermore, skeletal abnormalities such as bending distortion of long bones were more common among the British samples compared to St Mary’s samples. This indicates that, although the early industrialisation produced metabolic stresses, change of the environment through colonisation of new continents altered the distribution of metabolic deficiencies.

The new colony of South Australia, with its natural resources and Mediterranean climate could have offered many early British migrant the hope of a better life. However, unpreparedness of the Government of the new colony for settlement of the early migrants, and the refusal of the British Government to bear the cost of the establishment of the new infrastructure, led to an economic recession and mass unemployment. These caused marked hardship to the migrants. Subsequently, the timely action of the third governor of the colony helped the South Australian economy to recover. The pattern of burials at the “free ground” area of St Mary’s Cemetery from 1847 to 1927 showed the evolution of the economic status of some migrants who had settled at St Marys-on-the-Sturt. Furthermore, this pattern of burials in the St Mary’s Cemetery was a good reflection of the wider economic development of the colony.

*The authors have no conflicts of interest to declare*.

*This research received no specific grant from any funding agency, commercial entity or not-for-profit organization*.

*The data that support the findings of this study are available in the supporting information or from the corresponding author upon request*.

### Professional and Funding Acknowledgements

The authors acknowledge The University of Adelaide including the Faculty of Health and Medical Sciences Honours and PhD Scholarships for AG. Fr. William Deng, St Mary’s Anglican Church and the Department of Archaeology, Flinders University, for allowed access to the St Mary’s skeletal sample. The authors acknowledge the facilities, and the scientific and technical assistance, of the Australian Microscopy & Microanalysis Research Facility at the Adelaide Microscopy, University of Adelaide, especially Ruth Williams for her assistance with micro-CT scanning and post processing software.

## Data Availability

The data that support the findings of this study are available in the supporting information or from the corresponding author upon reasonable request.

## Notes

### Competing Interest Statement

The authors have declared no competing interest.

### Author Declarations

Office of Research Ethics, Compliance and Integrity Research Services, The University of Adelaide, South Australia.

